# The effectiveness of psychological interventions for anxiety in the perinatal period: A systematic review and meta-analysis

**DOI:** 10.1101/2022.01.14.22269296

**Authors:** Natalie Clinkscales, Lisa Golds, Katherine Berlouis, Angus MacBeth

## Abstract

**Background:** Anxiety disorders are a relatively common occurring mental health issue during pregnancy and the perinatal period. There is evidence that untreated perinatal anxiety is a risk factor for adverse outcomes for mother and infant. Despite their potential acceptability to users, psychological interventions research for this population is still in its infancy. This systematic review and meta-analysis aimed to comprehensively evaluate the evidence of the effectiveness of psychological interventions for reducing perinatal anxiety.

**Method:** This review followed the Preferred Reporting Items for Systematic Reviews and Meta-Analyses (PRISMA) guidelines. Databases searched included EMBASE, MEDLINE, PsychINFO, MIDIRS, CINAHL and the Cochrane Library. Search terms included: Psychological Therapy, Perinatal Period, Antenatal, Postnatal, Anxiety, Obsessive Compulsive Disorder and Phobia.

**Results:** The search strategy identified 2,025 studies. A total of 21 studies published between 2004 and 2021 fulfilled inclusion criteria. Of those, 17 were included in the meta-analysis. Overall results indicated that psychological interventions were more effective than control conditions in reducing symptoms of perinatal anxiety with a medium post treatment effect size. Significant effect sizes were also identified for online, face-to-face, group and guided self-help treatment modalities.

**Limitations:** A small sample of studies are represented and limited to articles published in English. The review was unable to draw specific conclusions about what works (i.e. therapeutic modality/delivery) for whom (i.e. specific diagnoses) due to purposefully broad inclusion criteria. The longer-term effects of psychological interventions for perinatal anxiety and infant outcomes could not be established.

**Conclusions:** This review demonstrates that psychological interventions are effective in reducing symptoms of both anxiety and comorbid anxiety and depression in the antenatal and postnatal periods. The results also demonstrate the efficacy of delivering such interventions in multiple settings, including online, and in group format. Further research is required to optimise treatment delivery to individual needs.

## 1. Introduction

The perinatal period represents a time of increased risk for the development of mental health problems (Biaggi et al., 2016; O’Hara and Wisner, 2014) and exacerbation of pre-existing conditions (Higgins et al., 2018). There is growing body of literature exploring the impacts of anxiety in the perinatal period, from pregnancy to one-year post childbirth (Loughnan et al., 2018). Prevalence rates for self-reported anxiety symptoms are 24.6% during the third trimester of pregnancy and 15% during the postnatal period (Dennis et al., 2017); and the overall prevalence of diagnosed anxiety disorder during pregnancy is 15.2% during pregnancy and 9.9% postnatally (Dennis et al., 2017). Evidence also suggests that one in 10 women will experience comorbid anxiety and depression during pregnancy and one in 12 during the postnatal period (Falah-Hassani et al., 2017). An extensive range of anxiety disorders are prevalent in the perinatal period (O’Hara and Wisner, 2014). Leach et al. (2017) with reported rates of generalised anxiety disorder (GAD) ranging from 1-11%, specific phobia ranging from 7-20%, panic disorder (PD) ranging from 1-8% and agoraphobia ranging from 1-17%. Rates of obsessive compulsive disorder (OCD) are more prevalent in the postnatal period (4-9%) than in the general population (1.2%; McGuinness et al., 2011). However, despite the evidence suggesting that prevalence rates for perinatal anxiety are similar to, if not greater than, that of perinatal depression, until recently, research aimed at understanding and treating anxiety in this period has been largely neglected (Bauer et al., 2016; Buist et al., 2011Loughnan et al., 2018; Maguire et al., 2018).

Unrecognised and untreated perinatal anxiety has significant consequences for women, their infants and wider family (Maguire et al., 2018), including a higher likelihood of developing postpartum depression, negative effects on the mother-infant attachment, more risk of obstetric complications and adverse outcomes for fetal and infant development (Dunkel Schetter and Tanner, 2012; Glasheen et al., 2010; Glover, 2014; Milgrom et al., 2008). These impacts are a major concern for clinical and public health (Blackmore et al., 2016; Dennis et al., 2017), alongside the economic cost of untreated perinatal anxiety and depression (Bauer et al., 2016).

In comparison to that of depression, the literature on the effective treatment and clinical management of perinatal anxiety remains limited including psychological interventions (Loughnan et al., 2018; Marchesi et al., 2016). However, psychological interventions may be preferable to women in this period due to concerns regarding the impact on the fetus and on breast feeding, relating to pharmacological interventions (Green et al., 2020; Loughnan et al., 2018; Taylor et al., 2016).

Support for the utilisation of CBT for the treatment of perinatal anxiety was found in a meta-analysis of 13 studies examining its efficacy (Maguire et al., 2018). They reported large within groups effect sizes from pre-post treatment (*d* = 0.81) and from pre-treatment to follow up (*d* = 0.82) indicating that CBT is an effective treatment for perinatal anxiety (Maguire et al., 2018). However, smaller effect sizes were reported for between groups analyses (*d* = 0.49) meaning that CBT may not be more effective than non-active control conditions and further research is required to establish the superiority of CBT over alternative interventions (Maguire et al., 2018). This report was also subject to several methodological limitations including small sample sizes, high levels of heterogeneity, and overall low scores on quality assessment tools.

There is also tentative evidence for the use of mindfulness based interventions (MBIs), including mindfulness based cognitive therapy (MBCT) and mindfulness based stress reduction (MBSR), as an effective treatment for both anxiety and depression in this population (Shi and MacBeth, 2017; Woolhouse et al., 2014). It should be noted, however, that the majority of this research has been conducted with women during pregnancy and there is also significant methodological heterogeneity in this literature. In Shi and MacBeth (2017), 17 studies were identified, 16 of which were conducted in pregnancy and only one which involved participants in the first year after childbirth, suggesting gaps in the current literature.

Although psychological interventions can be delivered via individual, group, and internet delivered approaches (Bittner et al., 2014; Burger et al., 2020; Loughnan, Sie et al., 2019) there is a paucity of research directly comparing the modes of delivery. All seem to be acceptable to women based on rates of treatment adherence and patient feedback (Loughnan et al., 2018). However, internet-delivered interventions offer greater flexibility, which may improve access to treatment for women in this period when demands on their time are greater (Loughnan, Joubert et al., 2019). In a recent review, Loughnan, Joubert et al. (2019) identified only seven papers exploring the use of these interventions for anxiety and depression in the perinatal period. Of these papers none were targeted interventions for specific anxiety disorders or comorbid anxiety and depression. However, tentative conclusions were drawn in terms of the utility of these interventions for perinatal anxiety.

The current review builds on previous evidence evaluating the effectiveness of psychological interventions for reducing anxiety in the perinatal period. We focus specifically on studies where perinatal anxiety, or comorbid anxiety and depression were the primary intervention targets, regardless of therapeutic modality or mode of delivery. Unlike previous reviews (e.g. Maguire et al., 2018), studies of interventions for perinatal depression where anxiety is a secondary outcome were excluded. Specifically we asked:

- Are psychological interventions associated with reductions in anxiety during the perinatal period?
- Are psychological interventions for perinatal anxiety associated with improvement on secondary outcomes, i.e. improvement in general wellbeing, mother-infant attachment etc?
- Which modality of psychological interventions are most beneficial? I
- Is there a difference between interventions to reduce anxiety offered in pregnancy versus postpartum?
- What are the methodological sources of bias in the literature?

## 2. Method

### 2.1 Search Strategy

The systematic review search was conducted using PRISMA criteria (Page et al., 2021). Studies were identified by searching the electronic databases EMBASE, MEDLINE, PsychINFO, MIDIRS, CINAHL and the Cochrane Library. The following search terms were developed and combined using MESH terms and key words and adapted for use with each database: Psychological Therapy or Psychological Intervention or Psychotherapy or Cognitive Behavioural Therapy or Cognitive Therapy or Mindfulness or Mindfulness Based Cognitive Therapy or Mindfulness Based Approaches or Mindfulness Based Stress Reduction or Psychodynamic Psychotherapy or Group Psychotherapy or Interpersonal Psychotherapy or Psychological Treatment or Anxiety Management or Acceptance and Commitment Therapy or Compassion Focused Therapy AND Perinatal Care or Perinatal or Perinatal Period or Antenatal or Postnatal or Postnatal Care or Postpartum or Postpartum Period or Maternal or Pregnancy AND Anxiety or Obsessive Compulsive Disorder or Obsessions or Obsessive Behaviour or Compulsions or Compulsive Behaviour or Panic Disorder or Generalised Anxiety Disorder or Phobia or Fear of Childbirth or Tokophobia or Childbirth Trauma or Birth Trauma or Post-Traumatic Stress Disorder. Searches were conducted in August 2021. The initial search returned 2,025 articles, After abstract searching the full text of the remaining 140 articles were reviewed against the inclusion and exclusion criteria, with 21 studies meeting criteria of which 17 were included in the meta-analysis. This process is outlined in the PRISMA flowchart below (Fig. 1). Identified peer-reviewed studies were published between 2004 and 2021.

**Figure 1.**
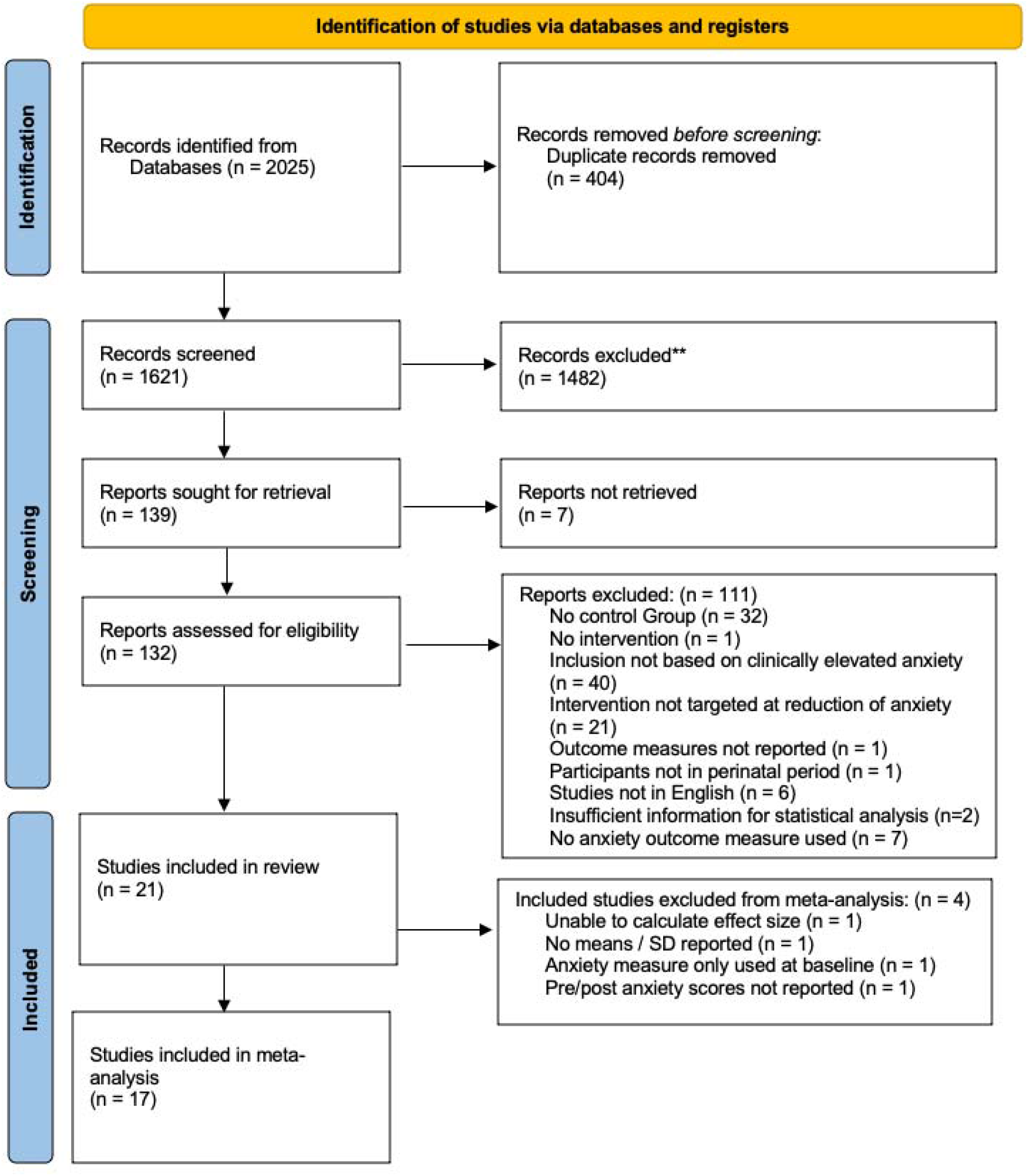
PRISMA Flowchart.

### 2.2 Inclusion and Exclusion Criteria

Studies were included if:

- They investigated the effectiveness of psychological interventions for treating anxiety in the perinatal period
- The reduction of anxiety or comorbid anxiety was a primary target of the intervention *or* where a reduction in other symptoms (e.g. depression) was targeted, the study was only included if participants were screened for anxiety as an inclusion criterion
- There was a treatment and control group
- Participants were women, over the age of 16, and pregnant or postpartum (up to one-year post-birth)

Participants anxiety was based on valid self-report measures or a clinical diagnosis of an anxiety disorder.

We women with a diagnosis of OCD, as although this has been reclassified in the US Diagnostic and Statistical Manual of Mental Disorders, fifth edition (DSM-V; American Psychiatric Association (APA), 2013) there remains a close relationship between OCD, anxiety disorders and other related disorders (APA, 2013). In addition, the psychological treatment interventions for OCD are similar to those offered for anxiety disorders and therefore treatment in the perinatal period is likely to overlap (Marchesi et al., 2016).

Studies were excluded if:

- The design was single case, case series or review
- They were not written in English (due to feasibility issues in accessing translations).

### 2.3 Data Extraction

#### 2.3.1 Demographics

A tailored proforma was developed to extract all relevant information from the full text of each eligible paper including: Citation, Country, Participants Characteristics, Sample Size, Age, Gestational Age at Baseline (weeks) or Infant Age, Study Design, Perinatal Period (Antenatal/Postnatal), Intervention, Clinician, Comparison Group, Diagnosis (Dx), Outcome Measure(s) and Assessment Time Point.

#### 2.3.2 Meta-analytic Model

Analyses were conducted in RStudio (RStudio Version 1.2.5033) using the ‘metafor’ (Viechtbauer, 2010) and ‘meta’ (Schwarzer, 2007) packages. It was assumed that the included studies would have a high degree of variability due to high methodological heterogeneity between studies, for example; therapeutic modality used (e.g. CBT vs Mindfulness) and method of intervention (e.g. group vs online). Therefore, random effects analyses were conducted applying the inverse variance method (Deeks et al., 2001), using DerSimonian Laird estimators for between-study variance (DerSimonian and Laird, 1986). Effect sizes were converted into Cohens d. Publication bias was investigated using visual inspection of funnel plots, and Egger’s test for plot asymmetry (Egger et al., 1997). Influence analyses were run to investigate the impact of outliers and impact of missing data modelled using a trim and fill analysis (Duval and Tweedie, 2000). Heterogeneity estimates were reported using I-squared values with values of 0, 25, 50, and 75% indicating zero, low, moderate, and high heterogeneity, in turn (Higgins et al., 2003).

#### 2.3.3 Risk of Bias Assessment

Risk of bias in the included papers was assessed using the revised Cochrane risk-of-bias tool for randomised trials (RoB2; Higgins et al., 2019). Bias was considered across 5 domains:

1. bias arising from the randomisation process
2. bias due to deviations from intended interventions
3. bias due to missing outcome data
4. bias in measurement of the outcome
5. bias in selection of the reported result

The first and second authors both completed the RoB2 template (see Appendix 2) independently for each study, following this both reviewers discussed and agreed their bias ratings for each study. Intra-class correlation coefficients based on Landis and Koch’s heuristics (1977) were conducted to establish level of agreement between reviewers.

## 3. Results

### 3.1 Study Characteristics

Demographic information are displayed in Table 1. A total of 21 studies were identified as meeting inclusion criteria, representing a sample of n=1,712 women (treatment conditions, n=777; control conditions, n=935). All participants were aged between 18-42, with a mean age of 29.8 years in the treatment group and 30.5 years in the control group. Due to inconsistent reporting, it was not possible to calculate the mean gestational/infant age. Of the 21 studies, seven were conducted in Iran, four in Australia, two in Canada, two in the UK, two in the Netherlands, and one each in Germany, China, USA, and Sweden. The interventions utilised were CBT (n=12), MBIs (n=4), a mixed approach self-help (n=2), Applied Relaxation (n=1), Cognitive Analytic Therapy (n=1), and Internet-based Problem-Solving Treatment (n=1). The methods of intervention delivery comprised of group setting (n=10), online format (n=5), face to face individual therapy (n=2) and guided self-help (n=4). The number of treatment sessions offered varied from two to 14. Anxiety outcomes were measured using the Spielberger State-Trait Anxiety Inventory (STAI; n=5), Generalised Anxiety Disorder 7-item scale (GAD-7; n=3), Beck Anxiety Inventory (BAI; n=3), Pregnancy Related Anxiety Questionnaire (PRAQ; n=2), Hospital Anxiety and Depression Scale (HADS; n=2), State-Trait Inventory for Cognitive and Somatic Anxiety, Trait Version (STICSA; n=1), Wijma Delivery Expectancy/Experience Questionnaire (W-DEQ-A; n=1), and the Hamilton Anxiety Rating scale (HAM-A; n=1). To measure depression the following outcome measures were used the Edinburgh Postnatal Depression Scale (EPDS; n=9), Patient Health Questionnaire 9-item scale (PHQ-9; n=4), Hamilton Depression Rating scale (HAM-D; n=1), Center for Epidemiologic Studies Depression Scale (CES-D; n=1), and Beck Depression Inventory-II (BDI-II; n=1). Of the 21 studies, ten completed measures only pre and post intervention and 11 studies administered follow up measures post intervention at varying timepoints. Due to the substantial variation in follow up time points meta-analytic modelling was not performed.

**Table 1:**
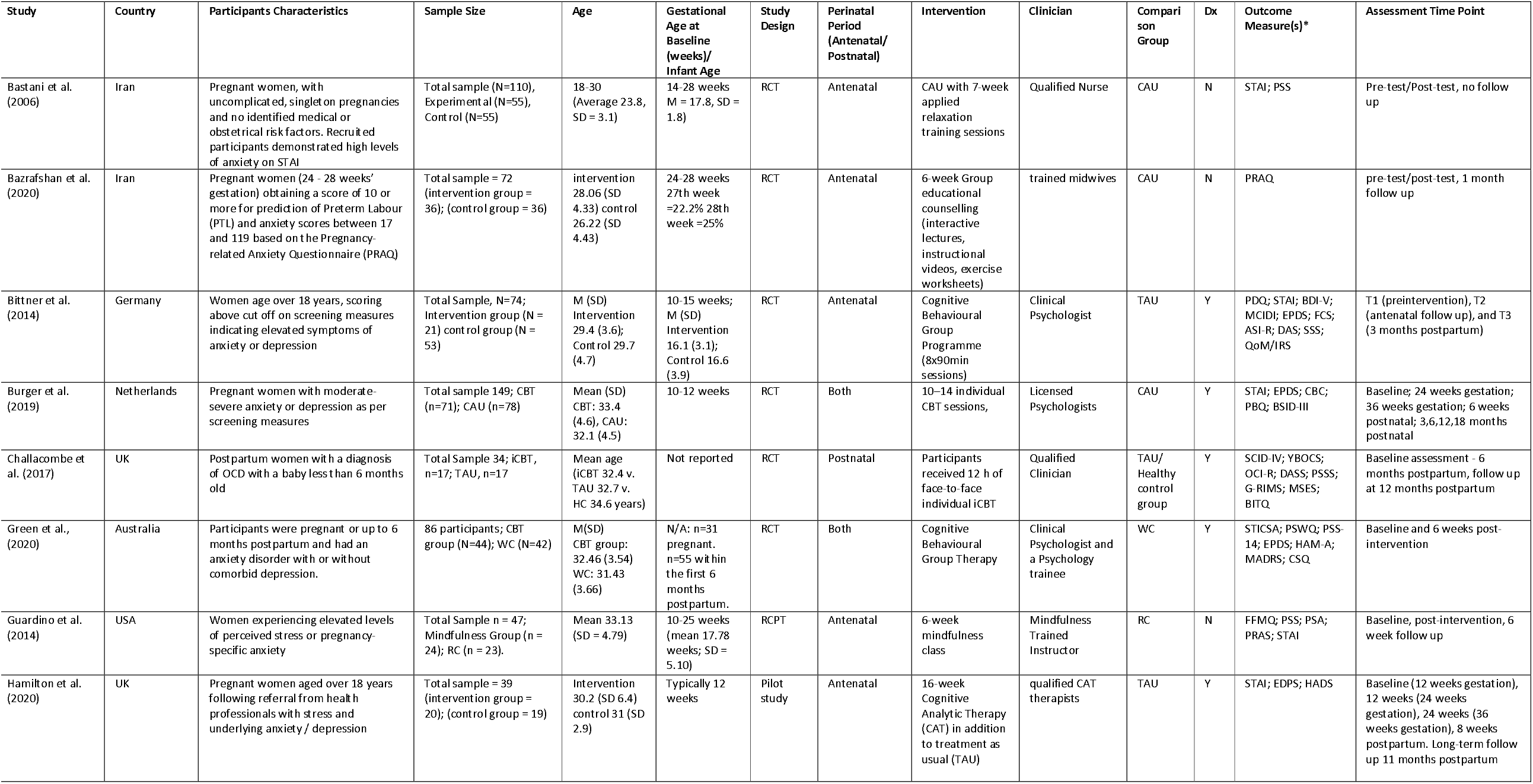
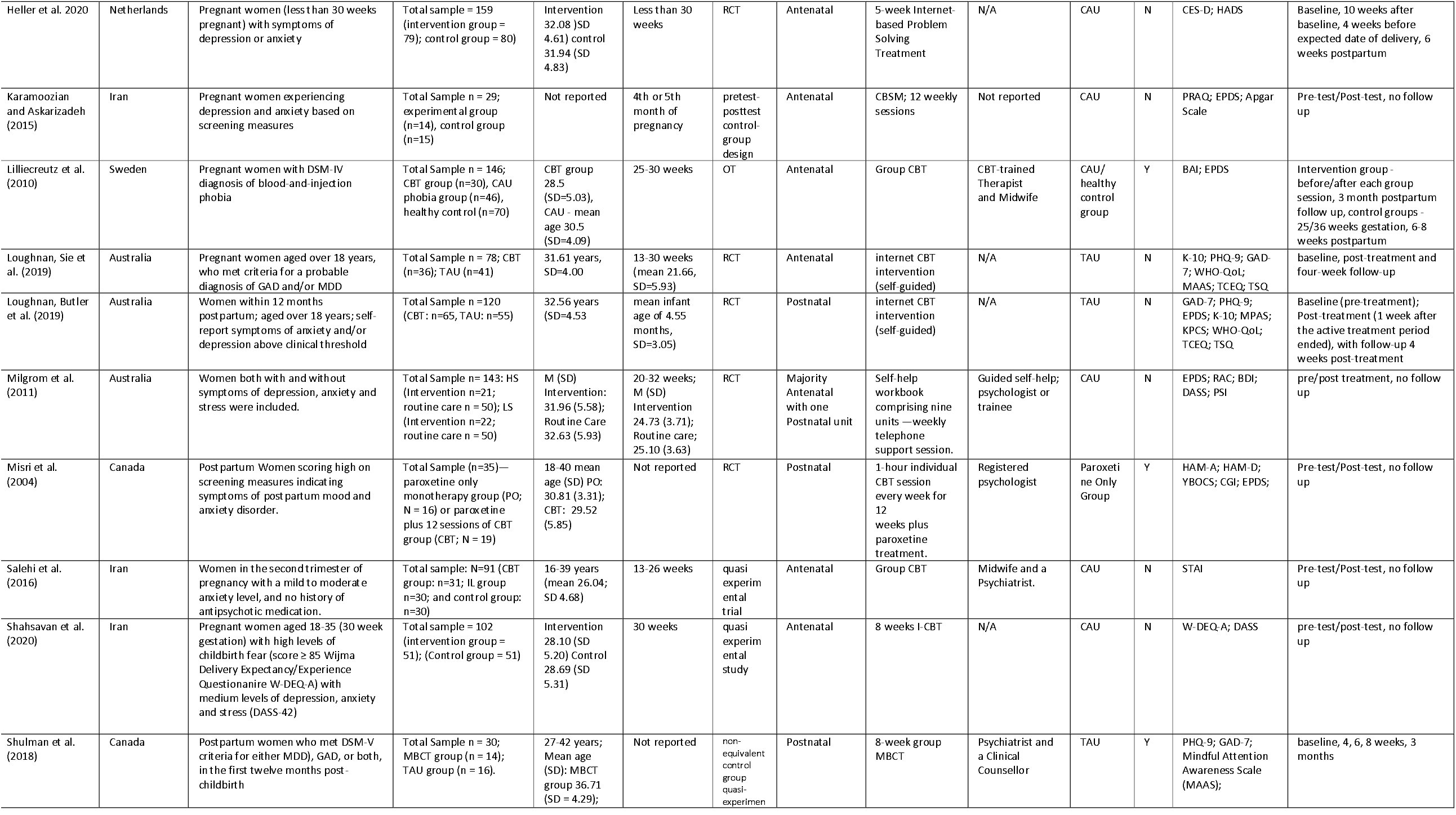
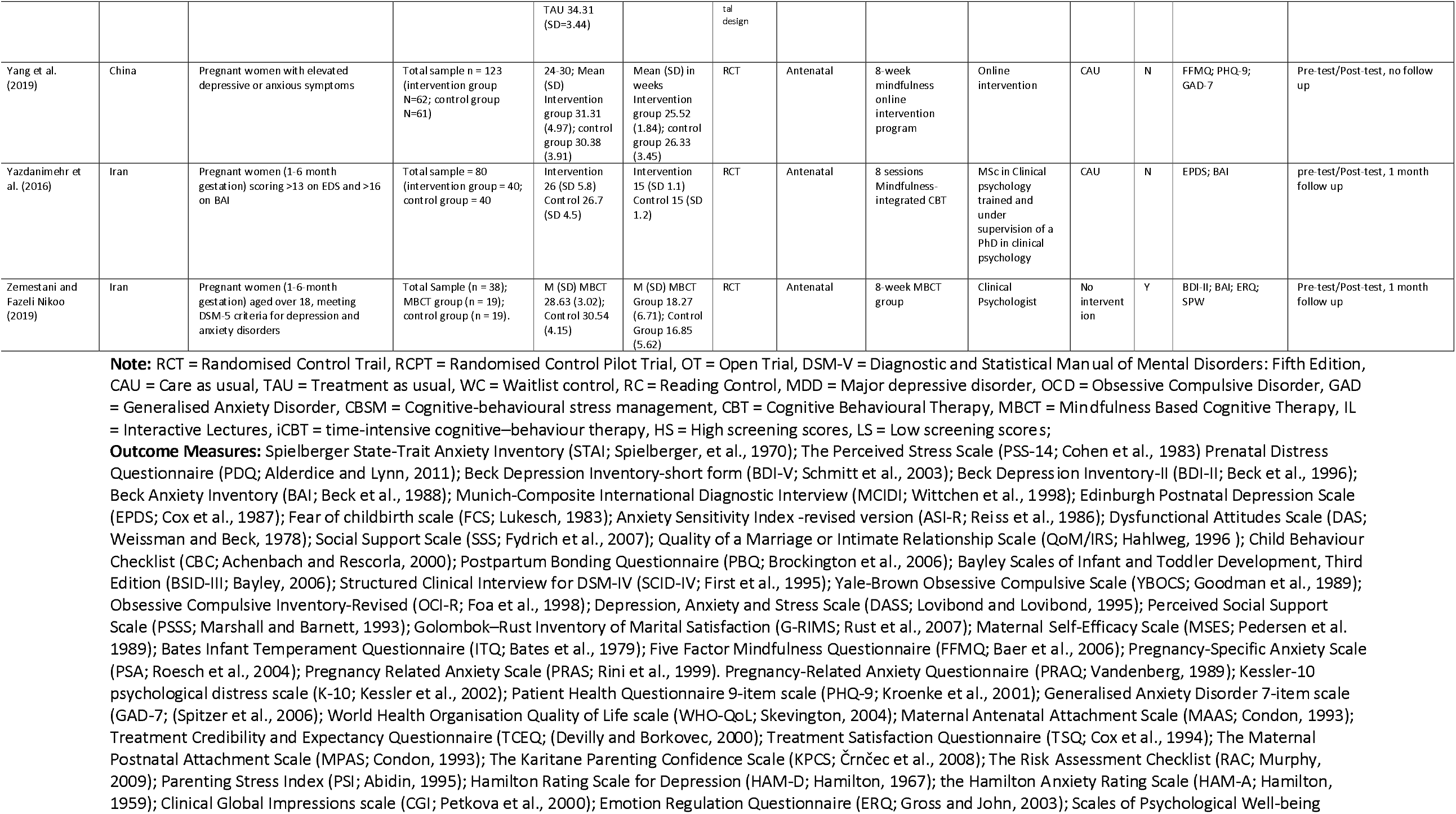
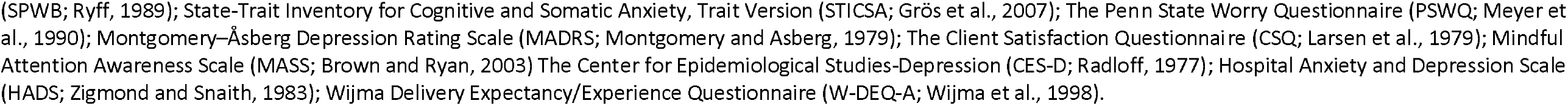
Study Demographic Characteristics.

### 3.2 Anxiety Scores Pre/Post Intervention

Results of the meta-analytic modelling of the effect of treatment on perinatal anxiety are displayed in Table 2. The main model (see forest plot, Fig. 2) reported a statistically significant effect size for the effect of intervention vs. control in reducing anxiety (*d* = -0.89; 95%CI = -1.35 to-0.43, *p*=0.002), indicating a medium effect with a broad but significant confidence interval. There was a high degree of heterogeneity between the studies (93.3%) indicating a considerable degree of between-study variance.

**Table 2:** Results of the meta-analytic modelling of the effect of treatment on perinatal anxiety.

| **Analysis (k)** | **Effect size (d) (95% CI)** | **Significance (p)** | **I<sup>2</sup> (%)** |
| --- | --- | --- | --- |
| **Anxiety Scores Pre/Post** | -0.89 (-1.35; -0.43) | **0.002** | 93.3 |
| *Anxiety Measures Subgroup* |  |  |  |
| *STAI (k=5)* | -0.19 (-0.60; 0.22) | N.S | 68.9 |
| **GAD-7 (k=3)** | -0.74 (-0.98; -0.48) | **<.05** | 3.1 |
| *BAI (k=3)* | -1.48 (-3.56; 0.59) | N.S | 96.8 |
| **PRAQ (k=2)** | -1.69 (-2.15; -1.23) | **<.05** | 0.0 |
| **Other (k=4)** | -1.13 (-2.61; 0.35) | N.S | 97.1 |
| *Type of Intervention Subgroup* |  |  |  |
| **Group (k=10)** | -0.94 (-1.49; -0.38) | **<.05** | 90.5 |
| *Self-guided (k=4)* | -1.27 (-2.64; 0.11) | N.S | 97.1 |
| *Individual (k=3)* | -0.0007 (-0.394; 0.393) | N.S | 34.1 |
| *Mode of Delivery Subgroup* |  |  |  |
| **Face to face (k=12)** | -0.74 (-1.26; -0.23) | **<.05** | 90.7 |
| **Online (k=5)** | -1.18 (-2.18; -0.17) | **<.05** | 96.1 |
| *Therapeutic Modality Subgroup* |  |  |  |
| **CBT (k=14)** | -0.80 (-1.31; -0.29) | **<.05** | 93.1 |
| *MBI (k=3)* | -1.28 (-2.78; 0.22) | N.S. | 94.6 |
| **Antenatal Treatment Only (k=13)** | -1.09 (-1.71; -0.47) | **0.0005** | 94.1 |
| **Effect of anxiety treatment on Depression scores (k=15)** | -0.97 (-1.49; -0.45) | **0.0002** | 93.5 |

Note: negative effect = reduction in anxiety, favouring treatment group

**Figure 2:**
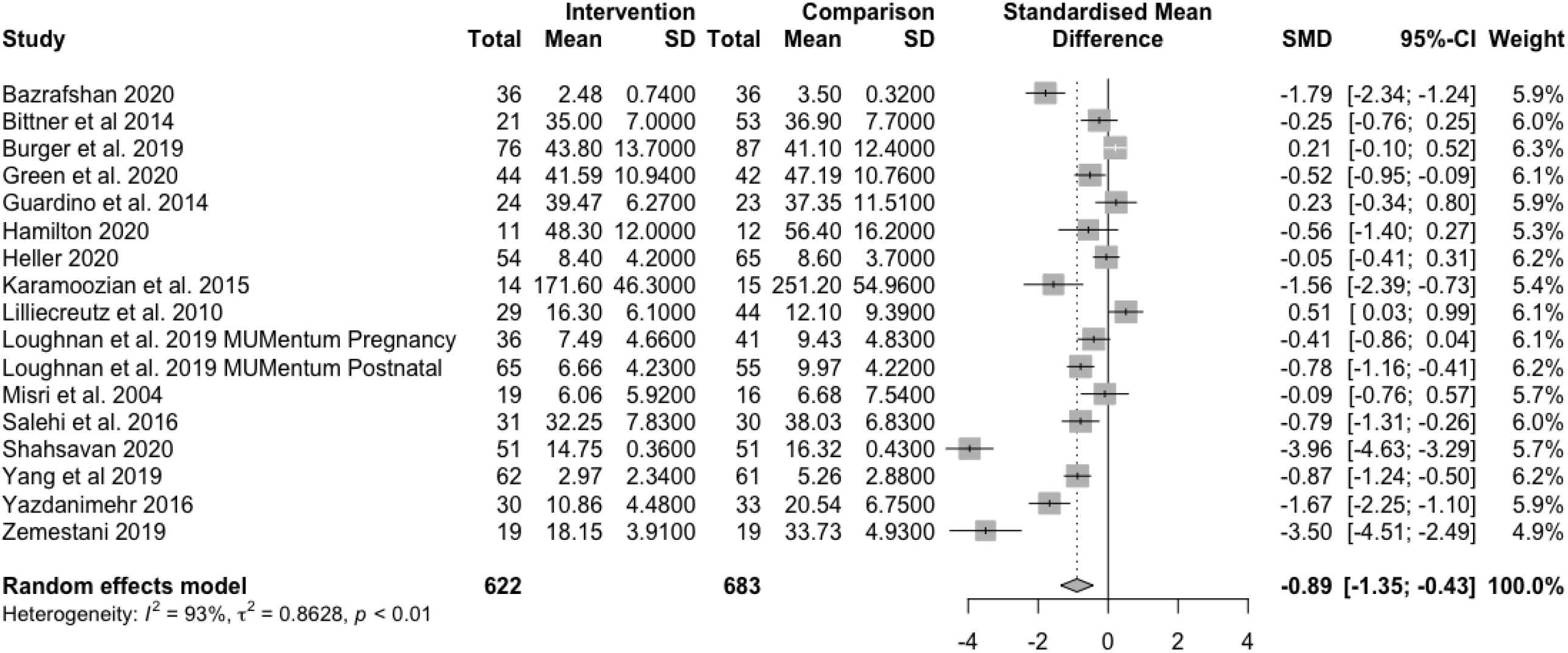
Forest Plot displaying the meta-analytic modelling of the effect of treatment on perinatal anxiety.

To investigate the potential impact of outliers, an influence analysis was also conducted. This analysis suggested that there was little effect of a leave one out analysis, with the exception of Zemestani and Fazeli Nikoo (2019) and Shahsavan et al. (2021). When these two studies were omitted, the overall effect size was reduced (*d*=-0.54) with reduced, but still significant confidence intervals (−0.87 to -0.21) as well as a reduction in heterogeneity (86.3%).

With regards to sensitivity analyses, visual inspection of the funnel plot suggested some asymmetry, confirmed using Egger’s test (B = -6.94, SE = 2.77, *p* = 0.02), indicative of potentially missing studies. The trim and fill method identified four missing studies. Included estimated missing effects in the analysis, suggested the effect for intervention vs. control in reducing anxiety would no longer be significant *k* = 21, *d* = -0.35 (95%CI = -0.89 to 0.19, *p* = 0.21).

### 3.3 Moderator Analyses

#### 3.3.1 Type of anxiety measure

In order to further explore these results subgroup analyses were conducted. With regard to the effect of different anxiety measures, effect sizes were significant for the GAD-7 (*d*=-0.74; 95%CI = -0.98 to -0.48) and the PRAQ (d=-1.69, 95%CI = -2.15 to -1.23). However, both estimates were based on small study numbers, indicating the effect was likely to be unstable. No other measures yielded significant differences between intervention and comparison. The test for subgroup differences was significant (*p*=0.0001) indicating that the type of anxiety measure used may impact the estimate for the effect of intervention on anxiety outcome in this context. Due to the small number of studies in each subgroup bias analyses were not conducted.

#### 3.3.2 Type of intervention

Next, moderator analyses examined the effect of type of intervention (group, individual or self-guided) on anxiety scores. For group interventions a significant medium effect size was found for the effect of intervention vs. control in reducing anxiety equivalent to *d* = -0.94 (95%CI = -1.49 to-0.38). This effect was based on ten studies with high heterogeneity (90.5%). No significant effects were found for individual or self-guided interventions vs. control for reducing anxiety. Overall, the test for subgroup difference between types of interventions was significant (*p*=0.01), suggesting type of intervention may impact upon overall anxiety reduction.

To account for outliers, the above analysis was re-run with the omission of the Zemestani and Fazeli Nikoo (2019) and Shasavan et al (2020) studies. For group interventions the overall effect size for the effect of intervention vs. control reduced to *d*=-0.71 (95% CI = -1.21 to -0.22), indicating a medium effect. Estimates for self-guided and individual interventions continued to be non-significant. The test for subgroup differences was non-significant (*p*=0.07) after removing the studies identified as outliers.

#### 3.3.3 Mode of delivery

For face-to-face interventions a significant medium effect size was found for the effect of face-to-face intervention vs. control in reducing anxiety equivalent to *d* = -0.74 (95%CI = -1.26 to -0.23). This effect was based on 12 studies with a high degree of heterogeneity (90.7%). A large effect was also found for online interventions vs. control in reducing anxiety (*d*=-1.18, 95% CI=-2.18 to -0.17). However, this effect was based on only five studies. Overall, the test for subgroup differences between mode of delivery was non-significant (*p*=0.45). However, due to difference in sample size between these two groups this result should be interpreted with caution.

Re-running the analyses with the omission of the outlying studies, Zemestani and Fazeli Nikoo (2019) and Shahsavan et al. (2021), indicated that for face-to-face interventions the overall effect size for the effect of face-to-face intervention vs. control reduced to *d*=-0.54 (95% CI = -1.00 to -0.08). The effect for online interventions was also reduced to a medium effect size of *d*=-0.53 (95% CI = -0.93 to -0.12). The test for subgroup differences remained non-significant (*p*=0.96). Again, due to small sample sizes, these results warrant further investigation.

#### 3.3.4 Therapeutic modalities

For CBT interventions a significant effect size was found for the effect of intervention vs. control in reducing anxiety equivalent to *d* = -0.80 (95%CI = -1.31 to -0.29). This effect was based on 14 studies with a high degree of heterogeneity (93.1%). However, we did not identify a significant effect of MBI interventions compared to controls. Overall, the test for subgroup differences between therapeutic modality was non-significant (*p*=0.55).

To account for outliers, the above analysis was re-run, omitting Zemestani and Fazeli Nikoo (2019) as well as Shahsavan et al. (2021). For CBT interventions, the overall effect size for the effect of intervention vs. control reduced to a medium effect size *d*=-0.56 (95% CI = -0.93 to -0.19).

#### 3.3.5 Effect of antenatal treatment

A subgroup analysis was also conducted for psychological interventions for anxiety offered in the antenatal period only. This model demonstrated an overall significant effect size for the effect of intervention vs. control in reducing anxiety in the antenatal period equivalent to *d* = -1.09 (95%CI = -1.71 to -0.47 *p*=0.0005), indicating a large effect with a broad but significant confidence interval. This result indicates that psychological interventions are effective for reducing anxiety in the antenatal period.

### 3.4 Depression Scores (Secondary Outcome) Pre/Post Intervention

Although the primary focus of the meta-analysis was on anxiety, the majority of included studies also measured depression, therefore a post-hoc meta-analysis was performed to explore the magnitude of effect of intervention on depressive symptoms. This model demonstrated an overall significant effect size for the effect of intervention vs. control in reducing depression equivalent to *d* = -0.97 (95%CI = -1.49 to-0.45, p=0.002), indicating a medium effect around a broad confidence interval. This result suggests that the psychological interventions were effective in reducing depression scores. There was a high degree of heterogeneity between the studies (93.5%).

To investigate the impact of outliers an influence analysis was also conducted. This analysis suggested that there was little effect of a leave one out analysis, with the exception of two studies; Zemestani and Fazeli Nikoo (2019) and Shahsavan et al. (2021). When omitted, the overall effect size was reduced to a small effect (*d*=-0.52) with confidence intervals between -0.86 and -0.19. The study heterogeneity also dropped to 83.5%. A sensitivity analysis was also conducted to explore the influence of bias. A visual inspection of the funnel plot suggested no asymmetry, confirmed using Egger’s test (B = -6.27, SE = 2.76, *p* = 0.04). The trim and fill method was applied and identified no missing studies.

### 3.5 Methodological Risk of Bias

The RoB2 methodological risk of bias tool (Higgins et al., 2019) was completed for each study by the first and second authors independently before consensus agreement was established (see Table 3 for consensus agreement ratings). Intra-class correlation coefficients indicated a fair level of agreement between reviewers (ICC (1) = 0.087) based on Landis and Koch’s heuristics (1977).

**Table 3:**
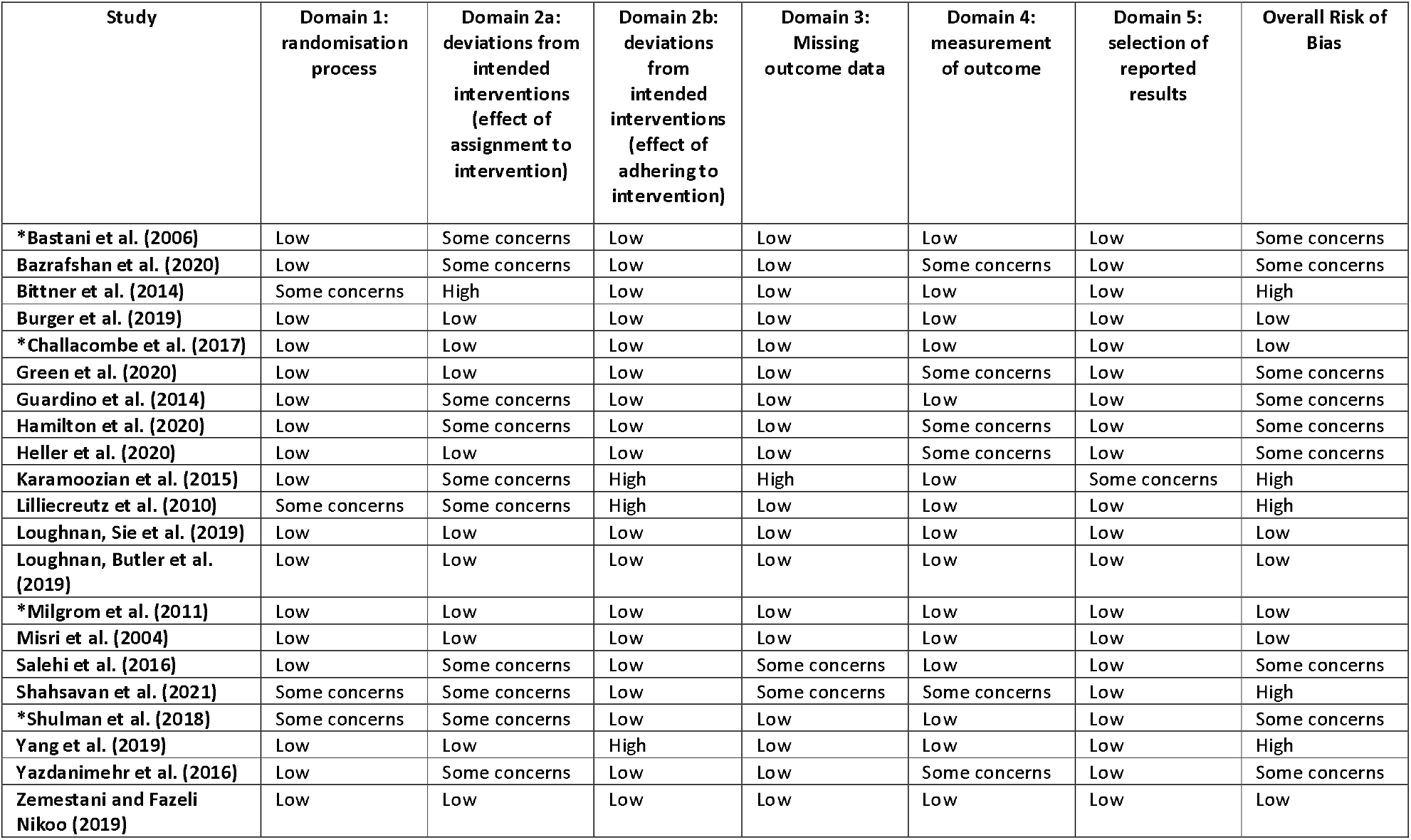
Consensus ratings between first and second reviewer on the RoB2. *Note: Studies excluded from meta-analysis

| Study | Domain 1:<br>randomisation<br>process | Domain 2a:<br>deviations from<br>intended<br>interventions<br>(effect of<br>assignment to<br>intervention) | Domain 2b:<br>deviations<br>from<br>intended<br>interventions<br>(effect of<br>adhering to<br>intervention) | Domain 3:<br>Missing<br>outcome data | Domain 4:<br>measurement<br>of outcome | Domain 5:<br>selection of<br>reported<br>results | Overall Risk of<br>Bias |
| --- | --- | --- | --- | --- | --- | --- | --- |
| *Bastani et al. (2006) | Low | Some concerns | Low | Low | Low | Low | Some concerns |
| Bazrafshan et al. (2020) | Low | Some concerns | Low | Low | Some concerns | Low | Some concerns |
| Bittner et al. (2014) | Some concerns | High | Low | Low | Low | Low | High |
| Burger et al. (2019) | Low | Low | Low | Low | Low | Low | Low |
| *Challacombe et al. (2017) | Low | Low | Low | Low | Low | Low | Low |
| Green et al. (2020) | Low | Low | Low | Low | Some concerns | Low | Some concerns |
| Guardino et al. (2014) | Low | Some concerns | Low | Low | Low | Low | Some concerns |
| Hamilton et al. (2020) | Low | Some concerns | Low | Low | Some concerns | Low | Some concerns |
| Heller et al. (2020) | Low | Low | Low | Low | Some concerns | Low | Some concerns |
| Karamoozian et al. (2015) | Low | Some concerns | High | High | Low | Some concerns | High |
| Lilliecreutz et al. (2010) | Some concerns | Some concerns | High | Low | Low | Low | High |
| Loughnan, Sie et al. (2019) | Low | Low | Low | Low | Low | Low | Low |
| Loughnan, Butler et al. (2019) | Low | Low | Low | Low | Low | Low | Low |
| *Milgrom et al. (2011) | Low | Low | Low | Low | Low | Low | Low |
| Misri et al. (2004) | Low | Low | Low | Low | Low | Low | Low |
| Salehi et al. (2016) | Low | Some concerns | Low | Some concerns | Low | Low | Some concerns |
| Shahsavan et al. (2021) | Some concerns | Some concerns | Low | Some concerns | Some concerns | Low | High |
| *Shulman et al. (2018) | Some concerns | Some concerns | Low | Low | Low | Low | Some concerns |
| Yang et al. (2019) | Low | Low | High | Low | Low | Low | High |
| Yazdanimehr et al. (2016) | Low | Some concerns | Low | Low | Some concerns | Low | Some concerns |
| Zemestani and Fazeli Nikoo (2019) | Low | Low | Low | Low | Low | Low | Low |

Overall, five studies were rated as having a high risk of bias, nine studies raised some concerns, and seven studies were rated low. For domain one, bias arising from the randomisation process, four studies raised some concerns, namely due to the lack of information regarding the actual randomisation method used. Domain two accounted for risk arising from deviations from intended interventions and is further broken down into two subcategories: a) effect of assignment to intervention and b) effect of adhering to intervention. In domain 2a, 10 studies raised some concerns, in all cases this related to the lack of intention to treat (ITT) analysis, or other appropriate analysis to estimate the effect of assignment to intervention. However, in these studies small sample sizes may mean there were fewer resources to complete ITT analysis. One study was rated as high in this area (Bittner et al., 2014), this was due to lack of appropriate analysis to estimate the effect of assignment to intervention, along with a large dropout rate, which may have had a substantial impact on results. For domain 2b, the majority of studies (n=17) delivered interventions to protocol and were, therefore, considered low risk of bias. However, three studies were rated as having a high risk of bias, this was primarily due to the lack of information regarding fidelity and adherence to treatment. For Yang et al. (2019) it was reported that adherence to treatment was low but appropriate analysis to estimate the effect of this was not conducted. In domain three, two studies were rated as some concerns, and one rated high, based on missing outcome data. For Karamoozian and Askarizadeh (2015) the risk was considered high due to inconsistent reporting. Overall, the measurement of outcome was appropriate (domain 4), however, six studies were flagged as having some concerns. Overall, the selection of reported results (domain 5) was appropriate for the majority of studies, indicating a low risk of bias. However, Karamoozian and Askarizadeh (2015) raised concerns due to lack of information regarding planned analysis

## 4. Discussion

The current meta-analysis aimed to systematically review and assess the evidence for the effectiveness of psychological interventions for anxiety in the perinatal period. Overall results indicated that psychological interventions were more effective than control conditions in reducing symptoms of perinatal anxiety with an estimate of a medium post-treatment effect size. Using a larger, and methodologically rigorous sample this result extends previous review evidence (Maguire et al., 2018). It is also in line with narrative reviews of the effectiveness of psychological interventions for perinatal anxiety (Loughnan et al., 2018; Marchesi et al., 2016) and consistent with results for treatment of anxiety in the general population (Newby et al., 2015; Watts et al., 2015).

The meta-analysis included a relatively small number of studies (n=17) with high levels of heterogeneity, therefore, results should be considered with a degree of caution. Moderator analyses also indicated a number of methodological aspects contributing to the effect size for anxiety reduction. Analyses indicated that no one outcome measure was superior in capturing anxiety scores, however, this result was based on small sample sizes. Of note, most studies used general measures of anxiety, with only three studies utilising a pregnancy specific anxiety measure as their primary outcome (Bazrafshan et al., 2020; Karamoozian and Askarizadeh, 2015; Shahsavan et al., 2021). It has been suggested in the literature that pregnancy-related anxiety may be a distinct concept which might uniquely predict obstetric outcomes and postpartum mood disorders (Blackmore et al., 2016). Furthermore, pregnancy-specific measures account for anxiety related to health of the baby, impact of previous miscarriages/obstetric complications and first time versus experienced mothers (Blackmore et al., 2016). These pregnancy specific fears and worries have not yet been explored in treatment research, and their potentially transient nature may account for some reductions in symptomology as the pregnancy progresses (Blackmore et al., 2016). Previous literature has also suggested that the use of general measures may hinder the accurate identification of anxiety specific to the perinatal period (Loughnan, Joubert et al., 2019). The current review did not compare the use of general measures against pregnancy specific measures. Future studies could utilise a combination of measures to make comparisons across general and pregnancy specific anxiety, with implications for targeting and tailoring of interventions.

In addition, medium size effects were found for group-based, but no effects were identified for individual and self-guided interventions. Previous preliminary evidence suggested that individually delivered CBT was more effective than group CBT for women with perinatal anxiety (Maguire et al., 2018) and similar conclusions have been drawn for the treatment of perinatal depression (Sockol, 2015). However, previous research exploring individual versus group CBT for anxiety disorders in children and young people found no significant differences, suggesting that both were equally effective in the treatment of anxiety (Wergeland et al., 2014). A larger body of evidence has evaluated the use of individual CBT in the general population than for group CBT (Whitfield, 2010) and so it is perhaps unsurprising that close scrutiny of group versus individual CBT for perinatal anxiety has not yet been conducted. However, the result of the current meta-analysis suggests that although group interventions are effective for this population, further evidence is required regarding individual or self-guided approaches.

For mode of delivery, both face to face (*d*=-0.74) and online (*d*=-1.17) interventions showed medium to large effect sizes suggesting similar benefits. This is the first review to compare internet delivered interventions for perinatal anxiety with face to face interventions (Loughnan, Butler et al., 2019). Although, a meta-analysis directly comparing the use of internet delivered CBT versus face to face CBT in the general adult population suggested the two formats are equally effective in the treatment of a range of psychiatric and somatic conditions, including various anxiety disorders (Carlbring et al., 2018). In addition, it has been highlighted that increasing the types of interventions and mode of delivery offered, such as group and internet based interventions, could have positive impacts on addressing treatment gaps and removing barriers to access (Kazdin, 2017). As such, exploring these factors for the perinatal population may be beneficial in determining the most effective and accessible treatments.

For therapeutic modality, the main therapeutic models utilised in the included papers were CBT and MBIs. Only CBT emerged as significantly associated with anxiety reduction, with no significant effect for MBI’s. However, we note that only 3 studies using MBI’s met our inclusion criteria. We also note that removal of outliers reduced the effect size for CBT (d=-0.56). There have been no previous reviews directly comparing therapeutic modality for the treatment of perinatal anxiety. Moreover, there is a paucity of research exploring the direct comparison between CBT and MBIs in the treatment of anxiety in the general adult population. However, in the treatment of depression, accounting for anxiety reduction as a secondary outcome, CBT and MBCT have demonstrated equal effectiveness (Manicavasgar et al., 2011). In addition, both CBT and MBSR have been shown to have equal effects in the treatment of anxiety and depression in individuals with autism spectrum conditions (Sizoo and Kuiper, 2017). The results of our analysis are in line with those conducted for these different populations, therefore, suggesting the applicability of CBT for treatment of anxiety, and comorbid anxiety and depression in the perinatal period. However, further research on MBI’s in this population is required.

Of the 17 studies included, only two were conducted in the postnatal period, while 13 were conducted in the antenatal period and two spanned both. Therefore, it was not possible to directly compare the effectiveness of interventions delivered antenatally versus postnatally. Subgroup analysis of the effect of intervention on symptoms of anxiety in the antenatal period found a medium effect, suggesting that psychological interventions are more effective for reducing anxiety symptoms in this period than control conditions. Existing literature highlights that antenatal anxiety is a strong predictor of postpartum mood disorders (such as depression; Loughnan, Joubert et al., 2019), therefore, this result supports the notion that delivering effective antenatal interventions may confer positive benefits for both mother and baby. This supports previous research which argues that early intervention antenatally can improve later outcomes for women and their infants (Thomas et al., 2014). However, further research is needed to establish the longer-term benefits of interventions offered antenatally and to evaluate treatments for postpartum anxiety. The delivery of interventions in the postpartum period may differ significantly from those delivered antenatally due to conflicting demands on new mothers (i.e. breastfeeding/childcare) and other factors, such as the impact of disrupted sleep.

This study also aimed to explore the secondary outcomes reported in the selected studies. A number of secondary outcomes were explored within the research, including perceived stress, worry, infant temperament, child behaviour, postpartum bonding and depression. The only secondary outcome consistently reported across studies was depression, thus limiting our statistical analyses of comorbid outcome to this variable only. It is perhaps unsurprising that depression was measured in the majority of included studies (n=15) given the high rates of comorbidity reported between anxiety and depression in this population (Falah-Hassani et al., 2017). The results indicated that psychological interventions were more effective at reducing symptoms of depression than control conditions, demonstrating a medium effect. This suggest potential cross- or transdiagnostic effects, whereby interventions targeted specifically at the treatment of anxiety could also have a positive effect on depression. This supports research that suggests transdiagnostic interventions targeting both symptoms of depression and anxiety, tailored to the perinatal period, may also be more beneficial than disorder specific interventions (Green et al., 2020; Loughnan, Butler et al., 2019). Further research is needed to explore the impact of interventions for anxiety on infant outcomes.

### 4.1 Limitations

We report a number of limitations. First, inclusion criteria for the review were broadly defined in order to capture the wide range of anxiety symptomology. However, this did not allow differentiation between specific anxiety disorder diagnoses. Therefore we weresunable to draw any conclusions about what works (i.e. therapeutic modality/delivery) for whom (i.e. specific diagnoses). This reflects the heterogeneity in the research evidence. Further research may consider whether interventions tailored to overall symptom reduction are sensitive enough to account for the broad range of presentations of anxiety. Future trials could utilise clinical diagnostic tools, however it is important to consider whether this risks excluding those who do not meet diagnostic criteria but still experience elevated anxiety and would still benefit from psychological intervention. Alternatively, future findings may point to the value of transdiagnostic interventions for perinatal anxiety (Loughnan, Butler et al., 2019).

There are a number of pregnancy and birth related factors that may contribute to increased anxiety in the perinatal period, including hyperemesis gravidarum (severe vomiting; McCormack et al., 2011), preeclampsia (Asghari et al., 2016), significant health concerns for the infant (Gorayeb et al., 2013), pregnancy loss (Markin and McCarthy, 2019) and birth trauma (Weinreb et al., 2018). Interventions aimed at treating anxiety within the context of these complex factors lay beyond the scope of the review. This review also did not include any studies exploring the use of psychological interventions to treat post-traumatic stress disorder (PTSD) in relation to birth trauma. PTSD has strong conceptual links with perinatal anxiety (Nieminen et al., 2016; Saisto et al., 2006) and so this exclusion may represent a significant limitation to the generalisability of the current findings and suggest a gap in the current literature which warrants further investigation.

Due to the small sample of represented studies, and the variance in time points of follow up data, we were unable to draw conclusions about the long-term effects of psychological interventions for this population. Robust evaluation of treatment effects at follow up are imperative in order to effectively demonstrate the value of these interventions. It is important that these effects are systematically evaluated as previous research has suggested that anxiety symptoms may decrease naturally in the first six months following birth (Vismara et al., 2016) and so this phenomenon needs to be accounted for when exploring intervention effects. The included papers also did not consistently explore long term impacts on the mother-infant relationship and child development. Future research should consider how these factors can be evaluated considering the current evidence to suggest untreated perinatal anxiety is detrimental to both mother and infant (Glover, 2014; Loughnan et al., 2018).

The papers included in this review were all published in the English language and in peer reviewed journals, this may have introduced potential bias towards positive findings. Risk of bias in studies also varied significantly, with factors indicating high risk of methodological bias in a number of studies. It may be important for future intervention research to demonstrate rigorous delivery and treatment adherence by ensuring treatments are delivered by more than one trained facilitator, that treatment adherence is appropriately assessed (i.e. through use of recordings) and that therapist competence is independently rated (Shi and MacBeth, 2017). These measures need to be transparently outlined so appropriate risk of bias assessments can be undertaken. However, limitations based on study quality reflect challenges faced across the spectrum of health-service based treatment research in perinatal and infant mental health due to the nature of the population. In addition, this paper, and the included research, refers to the treatment of perinatal anxiety in women and does not account for the experiences of trans or non-binary individuals who are anxious during pregnancy or the postnatal period. There is limited research exploring the experience of perinatal mental health for trans and non-binary people. However, there is an acknowledgement that there may be a heightened vulnerability to experiencing such difficulties due to a number of factors including prejudicial treatment by professionals, misgendering and history of trauma or adversity (Greenfield and Darwin, 2021). Further research is needed to understand the experiences of trans and non-binary people in the perinatal period to ensure they are not excluded from both research and healthcare.

### 4.2 Directions for future research

We suggest that further RCTs and implemention studies are required to compare psychological interventions for perinatal anxiety with alternative psychological interventions and modes of delivery, the current research relies too heavily on treatment as usual and waitlist controls. The current research base is also limited to the use of CBT and MBIs. It has been suggested that other treatment modalities such as Interpersonal Psychotherapy (IPT; Sockol, 2018), Acceptance and Commitment Therapy (ACT; Bonacquisti et al., 2017) and Compassion Focused Therapy (CFT; Cree, 2010) may have utility in the treatment of common perinatal mental health conditions, however, these have not received adequate research attention. The bias in the literature towards CBT may be reflective of current NICE guidelines which advocate its use in this population (National Institute of Health and Care Excellence, 2014). The current review did not explore the differences in clinician delivering the interventions, although this may be an important factor to consider as this may influence the intensity of the intervention (i.e. primary care vs. specialist services), the accessibility (i.e. availability of adequately trained therapists and supervisors), integration into existing care pathways and cost-effectiveness. This is especially relevant in low resource settings where access to specialist services, practitioners and financial resources are limited (Clarke et al., 2013).

### 4.3 Conclusion

Our review is the most comprehensive synthesis to date of the available evidence base of the effectiveness of psychological interventions for perinatal anxiety. This review demonstrates that psychological interventions, including CBT and MBI’s, are effective in reducing symptoms of both anxiety and comorbid anxiety and depression in the antenatal and postnatal periods. Given the negative consequences of untreated antenatal anxiety on postpartum outcomes, and women’s preferences for psychological over pharmacological interventions, the current review advocates for the availability and use of these interventions in pregnancy, as well as postnatally. In addition, the results support a wide variety of intervention methods and modes of delivery, including face to face, online, group and self-guided, suggesting that psychological interventions can be tailored to meet the individual and perinatal-specific needs of women in this period.

## Data Availability

All relevant data are contained in the manuscript and supplementary material.

